# Explicitly modeling genetic ancestry to improve polygenic prediction accuracy for height in a large, admixed cohort of US Latinos: Findings from HCHS/SOL

**DOI:** 10.1101/2025.03.21.25324423

**Authors:** Xin Wang, Tamar Sofer, Oleksandr Frei, Robert Kaplan, Krista M Perreira, Nora Franceschini, Humberto Parada, Laura Zhou, Ole A. Andreassen, Hector Gonzalez, Anders M. Dale, Iris J Broce

**Affiliations:** Department of Neurosciences, University of California, San Diego, San Diego, California, USA; Cardiovascular Institute, Beth Israel Deaconess Medical Center, Harvard Medical School, Boston, MA; Department of Biostatistics, Harvard T.H. Chan School of Public Health, Boston, MA; Department of Medicine, Brigham and Women’s Hospital, Boston, MA; Institute of Clinical Medicine, University of Oslo, Oslo, Norway; NORMENT, K.G. Jebsen Centre for Psychosis Research, Institute of Clinical Medicine, University of Oslo, Oslo, Norway; Department of Epidemiology and Population Health, Albert Einstein College of Medicine, Bronx, NY, USA; Public Health Sciences Division, Fred Hutchinson Cancer Center, Seattle, WA 98109, USA; Department of Public Policy, University of North Carolina at Chapel Hill, Chapel Hill, NC; Department of Epidemiology, University of North Carolina, Chapel Hill, NC, USA; Division of Epidemiology and Biostatistics, School of Public Health, San Diego State University, San Diego, CA, USA; Department of Biostatistics, University of North Carolina, Chapel Hill, NC, USA; Weill Institute for Neurosciences, Department of Neurology, University of California, San Francisco, UCSF, San Francisco, California, USA

## Abstract

Polygenic scores (PGS) offer moderate to high prediction accuracy for complex traits, but most are developed in European ancestry cohorts, reducing their performance in populations of other ancestries. This study aimed to improve standing height prediction, a heritable and ancestry-influenced trait, in an admixed Latino cohort (HCHS/SOL) by modeling ancestry using principal components (PCs) alongside PGS. SNPs were selected from a large European ancestry GWAS using various p-value thresholds, and weights were trained using traditional and penalized regression in the UK Biobank (UKB). PGS with PCs were trained separately in HCHS/SOL and UKB. Compared to PGS alone, modeling PGS with PCs substantially improved height prediction in HCHS/SOL (R² increase of ∼0.1), while mild improvements were observed in UKB (R² increase of ∼0.01). These results underscore the importance of incorporating genetic ancestry into predictive models for admixed populations, particularly when the trait exhibits ancestry-specific associations.

## Main

Genome-wide association studies (GWAS) have identified common genetic variants that influence the susceptibility to complex traits and diseases. Polygenic risk models aggregate these small-to-moderate contributions to create powerful polygenic scores (PGS) that provide improved predictive accuracy for quantitative traits and disease risk relative to any individual genetic marker^1,2^. Because the majority of GWAS and subsequent polygenic prediction analyses have been conducted on cohorts predominantly of European ancestry, the generalizability of PGSs to non-European and admixed populations is reduced^3–6^. Studies within the Hispanic Community Health Study / Study of Latinos (HCHS/SOL) and others have shown that polygenic score performance depends on how closely the ancestry of the study population is to the group used to develop the score^3,7,8^. As sample sizes of non-European cohorts increase with more diverse and generalizable population, the potential for discovery and prediction will improve^9^. However, practical interim approaches are needed to improve accurate predictions across diverse ancestries, thereby mitigating disparities in research outcomes.

The predictive accuracy of polygenic scores (PGS) for a trait depends on its SNP heritability. Standing height serves as a prime example of a highly heritable, polygenic trait, with heritability based on twin studies of approximately 70-90%^10,11^ and SNP heritability of about 40-50%^8,12^. High SNP heritability indicates that a significant portion of the trait’s variation is due to additive effects of SNPs. The SNP heritability provides a quantitative estimate of the asymptotic prediction performance using linear prediction, as the sample size increases to infinity^5^. The difference between the higher twin heritability and the SNP heritability is commonly referred to as “Missing Heritability”, and has been hypothesized to be due to rare genetic variants, structural variants, epistatic effects, and/or gene-environment interactions^13,14^. Thus, while SNP heritability sets a limit on the accuracy of PGS predictions, and as sample sizes grow, these predictions may approach this theoretical maximum performance, factors such as rare variants and non-additive effects can hinder PGS from fully realizing its potential.

An important limitation of current polygenic prediction methods is their poor generalization performance across ancestries^3^. PGS often exhibit reduced accuracy in populations with ancestries that differ from those used in the training GWAS due to differences in allele frequencies, linkage disequilibrium (LD) patterns, and/or causal effect sizes^3,4^. Factors influencing differences in allele frequencies and LD patterns include genetic factors influenced by natural selection, historical events such as colonization and forced migration, voluntary migration, and mating strategies^15^. Genetic principal components (PCs) provide a straightforward approach to capturing genetic structure patterns across ancestries. Adjusting for PCs in GWAS is important to prevent false-positive associations lacking biological relevance and to ensure that ancestry information does not affect the weight of individual SNP effects. However, some evidence suggests that when applying PGS to a validation cohort with ancestry different from the training cohort or GWAS, explicitly modeling ancestry information within that validation cohort can be beneficial for improving prediction accuracy^16,17^. One study applied this approach and demonstrated improved PGS prediction in European subgroups, including those with Scandinavian, Southern European, and Ashkenazi Jewish ancestry, for three pigmentation-related traits: natural hair color, childhood and adolescent tanning ability, and basal cell carcinoma^16^. This strategy is thought to enhance prediction accuracy by accounting for genetic structure and compensating for variations in allele frequencies and linkage disequilibrium that might not be adequately represented in the training cohort^16^. However, the extent to which explicitly modeling ancestry improves polygenic prediction accuracy in non-European, admixed individuals—such as Latinos with varying proportions of Amerindian, European, and African ancestry—remains unclear.

To better understand this, we pursued two objectives: (1) We explored how incorporating PGS with PCs affects the accuracy of polygenic predictions for standing height compared to using PGS alone. Our analysis focused on a European descent training cohort from the UK Biobank (UKB) and a highly admixed Latino validation cohort, specifically the Hispanic Community Health Study/Study of Latinos (HCHS/SOL), which includes individuals with diverse ancestry proportions^18^. We used the latest GWAS summary statistics for height^8^ to select SNPs. Standing height was chosen as our trait of interest due to its high heritability and known association with ancestry. To validate our results and minimize overfitting, we employed cross-validation schemes and used R² to assess predictor efficacy, particularly the impact of explicitly modeling ancestry. (2) We then compared PGS development using Lasso and Ridge penalized regression methods to established PGS approaches. Penalized regression methods analyze all SNPs, while applying regularization to manage multicollinearity and reduce overfitting. Specifically, Lasso regression (L1 regularization) shrinks coefficients based on their absolute values, often reducing some coefficients to exactly zero. This property makes it effective for SNP selection by eliminating less relevant predictors. Ridge regression (L2 regularization) shrinks coefficients based on their squared values, controlling for large coefficients. These methods have demonstrated superior performance to conventional PGS approaches across diverse populations^19^.

## Results

### Participants

We used two main cohorts for our analysis: 407,844 participants of European ancestry from the UK Biobank (referred to as “UKB EUR”) and 11,858 participants from the HCHS/SOL^18^, all with available genotype and phenotype data. Demographics are shown in Table 1. Fig 1 illustrates the varying proportions of genetic ancestry from three continental regions for HCHS/SOL: European, Amerindian, and African (see Methods). Continental ancestry proportions were estimated using a model-based approach with ADMIXTURE software^20^, assuming the presence of these three ancestral populations (African, European, and Amerindian for k = 3)^21^. Participants were classified into “predominant ancestry groups” based on the maximum value among the continental ancestry proportions.

**Fig. 1.**
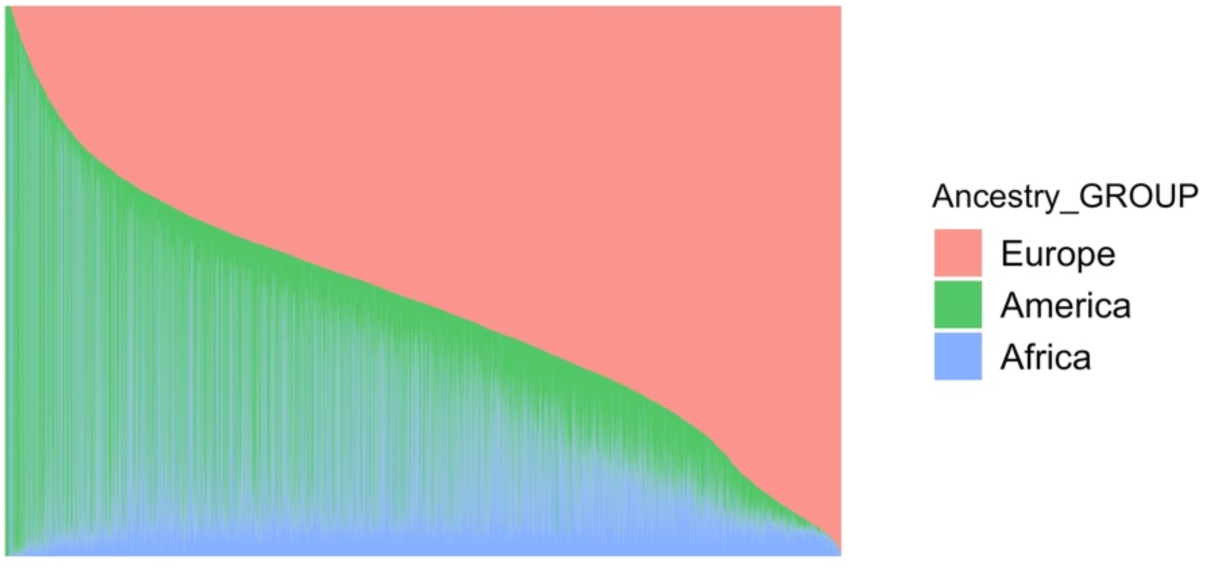
Varying proportions of genetic ancestry from three continental regions for HCHS/SOL participants.

**Table 1.** Demographics of participants.

|  | UKB EUR | UKB ALL | HCHS/SOL |
| --- | --- | --- | --- |
| N | 407,844 | 485,633 | 11,858 |
| Age (year) | 57.41(8.0) | 57.04 (8.09) | 46.08(13.82) |
| Sex, women N (%) | 220496 (54.1%) | 263440 (54.3%) | 6951 (58.6) |
| Standing height (cm) | 168.67(9.23) | 168.43(9.26) | 162.11(9.30) |

In supplementary analyses, we examined the entire UK Biobank sample (referred to as ’UKB ALL’), which includes an additional 77,789 participants from diverse ethnic backgrounds, including those of non-European origins. This was done to assess whether this inclusion improved the generalizability of our findings in HCHS/SOL.

### Mild improvement in prediction accuracy by modeling PCs with PGS compared to PGS alone when all training, validation, and GWAS cohorts are of European ancestry

We selected height-associated SNPs from the most recent and largest publicly available GWAS summary statistics for standing height derived from a cohort of European ancestry (excluding UK Biobank individuals)^8^ (Supplementary Fig 1). The number of SNPs used to construct PGS under different p-value thresholds is summarized in Supplementary Table 1 (see Methods). We split the UKB EUR cohort, using 90% of samples (n = 367,060) to develop the PGS with Lasso and Ridge penalized regression models, and allocated 10% of samples (n = 40,784) for validation. When modeling linear combination of PGS with PCs, we further divided the UKB EUR validation cohort, allocating 80% (n = 32,627) for training and 20% for validation (n = 8,157).

Using Lasso-derived PGS, when only the PGS for height was included in the prediction model (without PCs), prediction accuracy (R²) in UKB EUR validation cohort improved incrementally as more SNPs were included at higher p-value thresholds (Fig. 2). Specifically, the PGS R² was 0.27 when including SNPs meeting a 10^-16^ p-value threshold and improved to 0.387 when including SNPs meeting a 10^-2^ p-value threshold (Fig. 2). When PCs were added to the prediction model, R² increased slightly with each additional PC, showing a mild improvement of 0.01 in R² across all p-value thresholds (Fig. 2). The highest R^2^ reached to 0.396 at 10^-2^ p-value thresholds including 40 PCs. We also trained PGS using Ridge regression and observed similar trends as with Lasso regression (Supplementary Fig. 2). However, Lasso regression achieved the highest prediction accuracy of approximately 0.396 at lower p-value thresholds (0.01), whereas Ridge regression reached a similar R² at higher p-value thresholds (0.1).

**Fig. 2.**
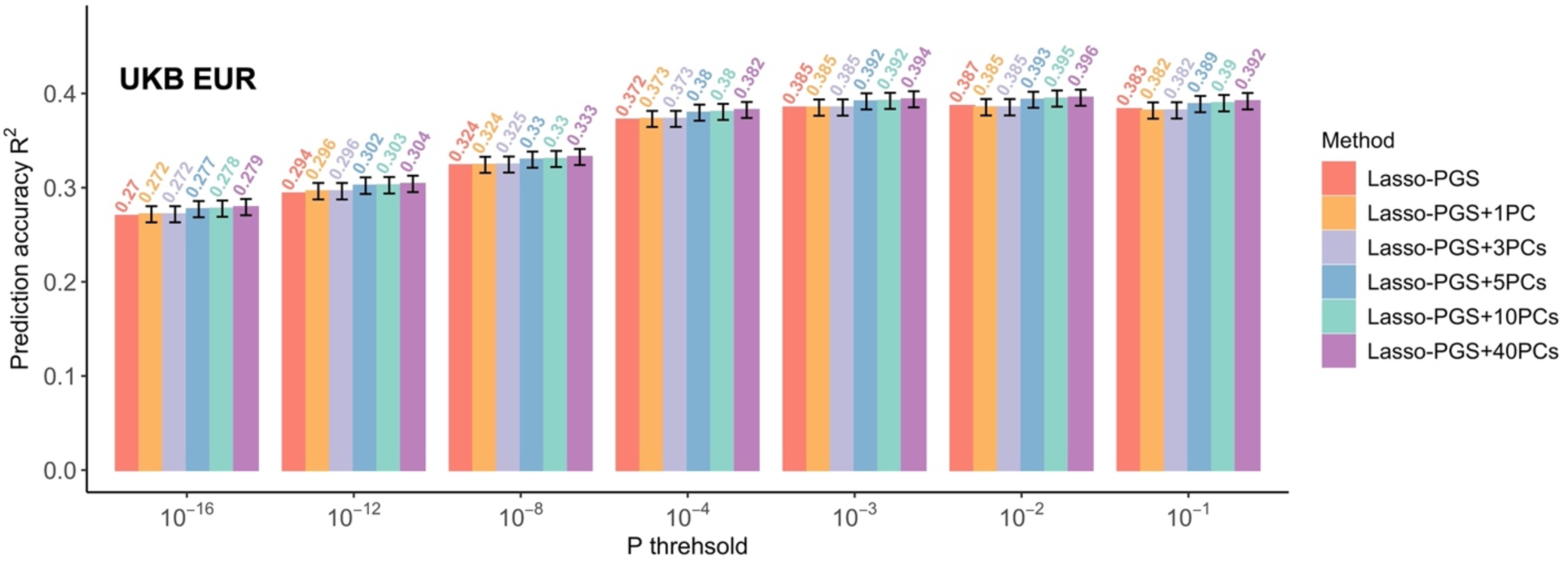
Comparison of predictive performance for height in the UK Biobank European (UKB EUR) validation cohort between using Polygenic Risk Scores (PGS) alone and a combination of PGS and Principal Components (PCs).

### Substantial improvement in prediction accuracy by modeling PCs with PGS compared to PGS alone when the validation cohort consists of admixed Latino individuals and training and GWAS cohorts were of European ancestry

For these analyses, we selected height-associated SNPs from the full EUR GWAS summary statistics for standing height (i.e., including UK Biobank), since HCHS/SOL was not part of the original GWAS discovery sample (see Methods) (Supplementary Fig. 1). The number of SNPs used to construct PGS under different p-value thresholds is summarized in Supplementary Table 1. We developed the PGS using the entire UKB EUR through Lasso and Ridge penalized regression models (same as above). We then applied the SNP weights derived from this model to the entire HCHS/SOL cohort. For modeling the PGS with PCs, we divided the HCHS/SOL cohort into a training set (80%, n = 9,486) and a validation set (20%, n = 2,372).

Using Lasso-based PGS, when only the PGS for height was included in the prediction model, the prediction accuracy (R²) varied across different p-value thresholds. The lowest R² (0.182) was observed when the fewest SNPs were incorporated at a p-value threshold of 10^-16^, while the highest R² (0.228) was observed at a p-value threshold of 10^-4^ (Fig. 3). Including PCs in the prediction model dramatically boosted the accuracy of height prediction, improving the R² by roughly 10% across all p-value thresholds. The highest prediction accuracy achieved was 0.318, observed at thresholds of 10^-4^ with the inclusion of 40 PCs. The greatest enhancement in predictive performance, ranging from 0.07 to 0.1, consistently occurred with the inclusion of the first 3 PCs across the different p-value thresholds (Fig. 3). Beyond the first 3 PCs, improvements were minimal, consistent with previous findings that suggest the first 2-3 PCs capture the majority of underlying variance^16^.

**Fig. 3.**
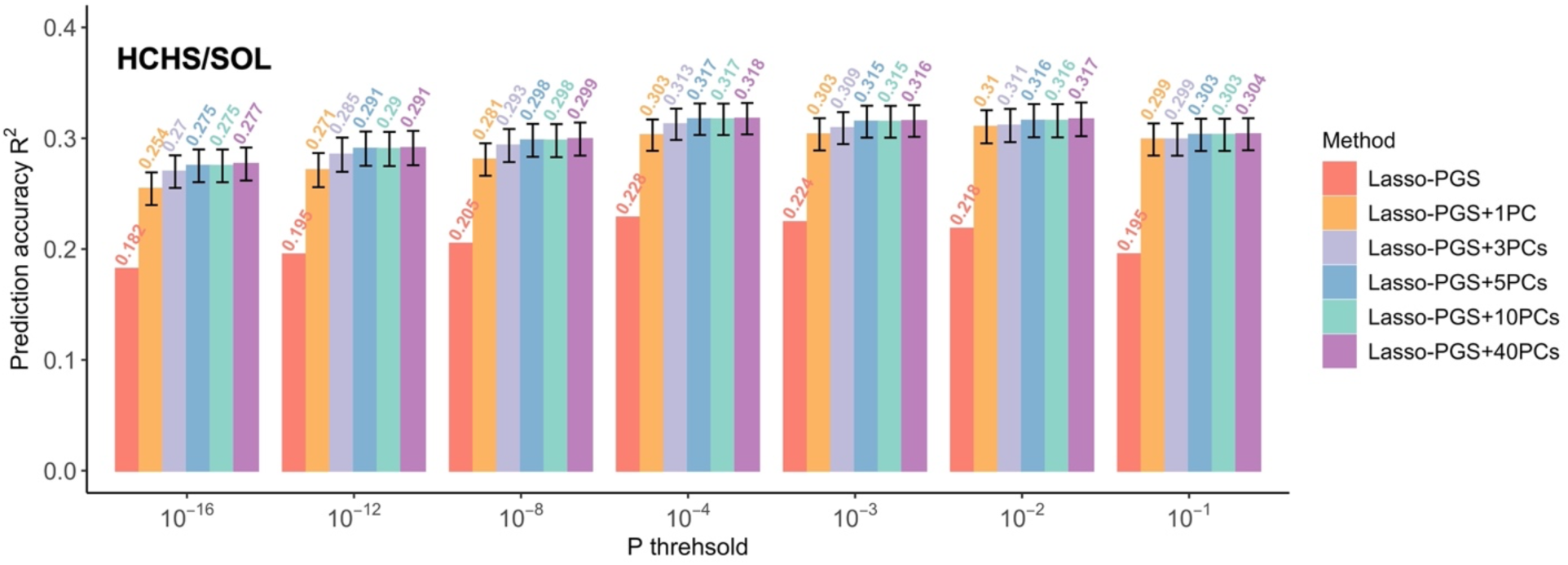
Comparison of predictive performance for height in the HCHS/SOL validation cohort using Polygenic Risk Scores (PGS) alone versus a combination of PGS and Principal Components (PCs). The SNP weights for the PGS were estimated using Lasso regression on the UK Biobank European (UKB EUR) training sample. When explicitly modeling PCs, we first split the HCHS/SOL validation cohort (n = 2,372) into 80/20 partitions. Linear regression models were fitted on the 80% training partition using standardized height as the outcome variable, and either PGS alone or a combination of PGS and PCs as predictors. The coefficients from these models were then applied to the 20% validation partition to predict height.

Additionally, we trained PGS using Ridge regression, and similar results were obtained showing that incorporating PCs significantly increased the prediction accuracy in HCHS/SOL (Supplementary Fig. 2).

### Across all three predominant ancestry groups in HCHS/SOL, the most significant improvement in predictive performance was achieved by including the first 3 PCs

We stratified the HCHS/SOL participants to three predominant ancestry groups based on their highest estimated ancestry proportion: European (n = 6,366), Amerindian (n = 2,764), and African (n = 733), and investigated the prediction accuracy within each group separately (Supplementary Fig. 3 and 4). In the European group, the highest prediction accuracy of the Lasso PGS was 0.255 at a p-value threshold of 10^-4^. Including PCs in the prediction model increased the R² by 0.01-0.03, with the highest R² reaching 0.276. For the Amerindian group, incorporating the PGS improved the prediction accuracy by about 0.08-0.12, with the highest prediction accuracy reaching approximately 0.32. In the African group, the prediction showed high variability, potentially due to the smaller sample size, but the inclusion of PCs increased the prediction accuracy by about 0.05-0.07, with the best prediction accuracy reaching 0.179. Across all three ancestry groups, the most significant improvement in predictive performance was consistently achieved by including the first 3 PCs (Supplementary Fig. 4).

### Increasing the training sample size by including individuals of varied ancestry enhanced out-of-sample prediction accuracy in HCHS/SOL

Following the same methods as above, we re-trained our PGS and PGS with PCs models using the UKB ALL as a training cohort and applied these weights to both the UKB ALL validation and HCHS/SOL cohorts (Supplementary Fig. 5). Similar to our results above, adding PCs slightly increased prediction accuracy in the UKB ALL validation cohort, with improvements ranging from 0.01 to 0.04. In contrast, the HCHS/SOL demonstrated a more substantial increase in prediction accuracy, ranging from 0.04 to 0.09 (Supplementary Fig. 6).

Lasso-based PGS models trained on UKB ALL consistently outperformed those trained on UKB EUR across both datasets. In UKB, the UKB ALL model exceeded the UKB EUR model by 0.01 in R², reaching a maximum R² of 0.409 (Supplementary Fig. 6A and Fig. 2). Similarly, in HCHS/SOL, the UKB ALL model outperformed the UKB EUR model by 0.01-0.02, both with and without PCs, achieving a highest prediction accuracy of 0.327 (Supplementary Fig. 6B and Fig. 3).

We also applied the PGS trained in the UKB ALL model to different predominant ancestry subgroups in HCHS/SOL. Across predominant ancestry groups, the model trained in UKB ALL also consistently outperform the models trained in UKB EUR by 0.01 (Supplementary Fig. 7 and Supplementary Fig. 4).

These findings are significant because they suggest that including diverse individuals in the training cohort can enhance out-of-sample predictions. This improvement may be due to the increased statistical power in determining SNP weights and the incorporation of PCs, which enhances model performance in cohorts with people from diverse ancestries.

## Discussion

This study evaluated how modeling genetic ancestry information, through principal components (PCs), alongside PGS impacts the prediction accuracy of standing height, with a focus on improving generalizability of PGS to non-European and admixed populations. We used the most recent and largest GWAS summary statistics for standing height to select SNPs and trained the PGS within the UKB using Lasso and Ridge penalized regression methods. We applied these PGSs to both the UKB and the HCHS/SOL, an admixed cohort of Latino individuals. We assessed prediction accuracy using the R² metric. Collectively, our results demonstrated that explicitly modeling ancestry information with PGS substantially improved height prediction performance in HCHS/SOL compared to using PGS alone. Additionally, including individuals of diverse ancestries in the training sample, rather than exclusively using a homogenous group of Europeans, further improved out-of-sample prediction accuracy in HCHS/SOL. These findings emphasize the need to incorporate ancestry information into PGS models and include diverse samples in the training cohort to improve generalizability to non-European and admixed populations. Additionally, our results suggest that continued data collection in diverse cohorts is essential for reducing disparities in research outcomes.

Previous studies have shown that explicitly modeling ancestry, even within European subgroups (Scandinavian, Southern European, and Ashkenazi Jewish ancestry), can enhance the predictive performance by R^2^ of 0.02 - 0.03 for ancestry associated traits, such as hair color, tanning ability, and basal cell carcinoma^16^. In our European UKB cohort, we observed slight prediction improvements (R^2^ of ∼0.01, depending on p-value thresholds) when modeling ancestry with PGS. The mild gain is likely due to the cohort’s pre-selection of “White British” individuals with PCs confirming European ancestry, minimizing ancestry-related genetic variation. However, when the training cohort and the validation cohort were from different ancestral backgrounds— such as applying PGS derived from a UKB EUR cohort to the HCHS/SOL —modeling ancestry information within HCHS/SOL alongside PGS substantially enhanced prediction accuracy (R^2^ of 0.07 to 0.1). Collectively these findings suggest that if you goal is to improve prediction, adding ancestry information can be beneficial, particularly when the training and validation cohorts differ in ancestry and the trait of interest is ancestry-dependent.

We found that the highest prediction accuracy for height in HCHS/SOL did not match the peak accuracy achieved in the UKB under any condition. In the original GWAS, Yengo and colleagues reported height prediction accuracy using traditional PGS approaches of R^2^ of about 0.40 - 0.44 in individuals from European ancestry and R^2^ of about 0.13 - 0.20 in a Latino/Hispanic group^8^. We applied this existing polygenic risk scores for height to HCHS/SOL, a independent Latino/Hispanic cohort from theirs, and observed similar results (R^2^= 0.218). We did not apply this existing PGS to our UKB EUR cohort because UKB data was included in the Yengo et al. training dataset, which would lead to overfitting, while HCHS/SOL was not included in the training set. Moreover, other groups have used less traditional PGS approaches, such as non-linear machine learning and report that that roughly 22% height variance is explained in their diverse Latino group^22^. Using Lasso and Ridge regression alone, we achieved prediction accuracy of about 0.19 – 0.23 in HCHS/SOL and 0.27 - 0.39 in UKB EUR (depending on p-value thresholds). Notably, our results in the UKB cohort did not reach the 44% accuracy observed in most recent Yengo et al GWAS, likely due to the lower sample size in our UKB training cohort (n = 367,060) compared to Yengo et al GWAS^8^ (n = 3.5M). While Lasso regression improved prediction accuracy in HCHS/SOL compared to other studies^6,8,22^, there remains significant potential to refine polygenic scores to improve accuracy in diverse populations, as current methods still perform better in European populations than in non-European groups.

Previous research within HCHS/SOL assessed the generalizability of polygenic scores (PGS) from European populations to Hispanics/Latinos.^6^ The study evaluated PGS performance using various strategies for SNP selection and weighting, including effect sizes estimated from large European GWAS and an admixed training population (HCHS/SOL), as well as from a fixed-effects meta-analysis combining both European and HCHS/SOL GWAS results. For height, PGS constructed with meta-analysis-derived effect sizes explained the highest proportion of variance (12%) in Hispanics/Latinos from the Women’s Health Initiative (WHI, n = 3,582).^6^ There are a couple difference between our study and this one. First, we utilized the most recent and largest height GWAS by Yengo et al. (2022) to select SNPs, which included a sample size of 3.5 million participants for SNP selection, whereas this study relied on the Wood et al. (2014) GWAS, which had a sample size of 253,288. The larger sample size in our study may enhance the statistical power and precision of existing effect size estimates, potentially leading to improved PGS performance in the Hispanic/Latino population. We also estimated effect sizes in the UK using a cohort that included both European and non-European participants, and we validated our results in HCHS/SOL, not WHI. We found that using Lasso and Ridge regression alone, we achieved a prediction accuracy (R²) of approximately 0.19–0.23 in HCHS/SOL and an R² of 0.218 when applying the most recent published Yengo GWAS PRS weights to HCHS/SOL. This is comparable to Yengo et al findings in Hispanic/Latino populations, which reported an R² of 0.13–0.20. Despite these differences, Grinde et al. found that selecting variants based on European GWASs generally performs well; however, for weight estimation, their approach was less optimal than including diverse individuals, a finding that aligns with our results.

The true novelty of our study lies in modeling PCs with PGS within our Latino cohort, which improved prediction accuracy by approximately 10% compared to PGS alone. Deriving PCs is relatively straightforward and computationally efficient. Further, when the training sample included individuals of diverse ancestries and we modeled PCs, prediction accuracy reached the highest in both UKB (0.409) and HCHS/SOL (0.327). The observed improvement may be attributed to both the larger sample size and the diverse ancestries in the training cohort. This finding highlights the importance of broader recruitment strategies that include diverse populations to enhance the generalizability of PGS.

This study has several limitations. First, the small sample size in the HCHS/SOL cohort, especially within different ancestral subgroups, limits the statistical power of our analyses in subgroup analyses within different predominant ancestry groups. Second, our analysis focused on height, since it is a highly heritable and well-studied trait. However, it is crucial to apply our approach to a broader range of heritable traits and diseases to assess its generalizability to health-related outcomes. Additionally, the generalizability of our findings may be limited by the specific methodologies used for PGS and the choice of prediction models, such as Lasso and Ridge regression. Lasso regression, while effective, is sensitive to missing values and may be less robust when replicating findings in other cohorts with varying degrees of missing data. To mitigate the impact of these limitations, we included SNPs that were shared across our GWAS, training, and validation cohorts. Also, while we did not comprehensively compare our results to existing PGSs, we included a comparison to the most recent height PGS from Yengo et al. as a reference point and discuss findings from previous studies. Another limitation is that our study focused on incorporating PCs to account for ancestry, which may not fully capture local ancestry effects. While our approach with PCs has demonstrated promise, it is possible that incorporating local ancestry effects could offer additional improvements in prediction accuracy by accounting for finer-scale genetic variations that PCs might overlook. Addressing these limitations in future research will be important for refining polygenic prediction models and enhancing their applicability across diverse populations and traits.

In summary, explicitly modeling ancestry information with PCs in PGS models and including diverse samples in the training cohort significantly improve the generalizability of PGS to non-European and admixed populations, with our study demonstrating that adding PCs increased prediction accuracy by approximately 10%. Despite this advancement, prediction accuracy for height in admixed populations, such as HCHS/SOL, remains below that achieved in a European ancestry cohort. Therefore, increasing recruitment efforts in diverse populations remains essential to enhance the generalizability and effectiveness of polygenic prediction models.

## Methods

### UK Biobank

UK Biobank is a large-scale prospective epidemiological study of over 500,000 individuals aged 40–69 years from the United Kingdom, which was established to investigate the genetic and non-genetic determinants of middle- and old-age diseases (https://www.ukbiobank.ac.uk/).

The UKB released genetic data for 487,409 individuals in 2018 (version 3). These samples were genotyped using either Affymetrix UK BiLEVE Axiom or Affymetrix UK Biobank Axiom arrays (Santa Clara, CA, USA), which includes over 800,000 genetic variants. The UK Biobank researchers implemented extensive quality control (QC) procedures on genotype data^23^ Imputation was performed centrally using combined data from the 1000 Genomes Project and the UK10K panel^24^, with SHAPEIT3 used for phasing and IMPUTE2 used for imputation^25^. Additionally, we excluded variants with a minor allele frequency (MAF) < 0.5% and multi-character allele codes. These QC criteria resulted in the inclusion of 10,188,578 SNPs. Forty PCs of genetic ancestry were supplied by the UK Biobank (Field 22009).

Standing height (cm) was supplied by UK Biobank (field 50). In this study, we included 485,633 individuals with height information and genotype data. Among them, 407,844 were Europeans who self-identified as ’White British” (Field 21000) and exhibited similar genetic ancestry based on PCA of their genotypes.

### The Hispanic Community Health Study/Study of Latinos (HCHS/SOL)

The HCHS/SOL^18,26,27^ is a population-based longitudinal cohort following Hispanic/Latino participants from four metropolitan areas: Bronx NY, Miami FL, Chicago IL, and San Diego CA, with 16,415 participants aged 18–74 years examined in the baseline visit. Participants were recruited through a two-stage sampling process. First, community block units were selected, and then households within those blocks were sampled. Some or all members of each selected household were invited to participate. 12,803 study participants consented to genetic studies^6,26^. Participants self-identified with six Hispanic/Latino background groups: Central American, South American, Mexican (Mainland groups, have high Amerindian genetic ancestry and low African ancestry), Cuban (high proportion of European ancestry, low African and Amerindian ancestry proportions) Dominican, and Puerto-Rican (Caribbean group, have low Amerindian ancestry, and high African ancestry proportions).^28^ In this study, we included 11,858 individuals who participated in HCHS/SOL study and were genotyped. All individuals provided written informed consent at their recruitment site.

### Genotyping

HCHS/SOL genetic data was assessed from blood during the baseline exam. Samples have been genotyped using three versions of the biobank SNP array offered by Illumina that is designed to capture the diversity of genetic backgrounds across the globe. The first batch of data was generated on the Multi-Ethnic Genotyping Array (MEGA) array, the first release of this SNP array. The second, third, and fourth batches were generated on the Expanded Multi-Ethnic Genotyping Array (MEGA Ex) array. All remaining data were generated on the Multi-Ethnic Global (MEG) BeadChip.^29–31^ Individuals who consented to genetic studies were genotyped using Illumina MEGA array, and a total of 11,928 samples and 985,405 genotyped variants passed quality control. For additional details about the DNA sampling see. ^29–32^

Genotype data were imputed to the Trans-Omics in Precision Medicine (TOPMed) freeze 5b reference panel as described before.^33^ Quality-control procedures in HCHS/SOL, including methods used to construct kinship matrix reflecting genetic relatedness between study participants (some of whom were sampled from the same household), have been described previously.^21^ Principal components analyses were performed using FlashPCA2.^34,35^ Global ancestry proportions measuring the proportion of the genome inherited from European, African, and Amerindian ancestors, and genetic principal components, were computed as previously reported^21^. Continental ancestry proportions were estimated using a model-based approach with ADMIXTURE software^20^, assuming the presence of three ancestral populations (African, European, and Amerindian for k = 3)^21^. We classified the participants into three ancestry groups based on the maximum value among the continental ancestry proportions.

In this study, we included 11,858 individuals who participated in HCHS/SOL study and were genotyped. All individuals provided written informed consent at their recruitment site.

### Summary statistics from published GWAS

Summary statistics were derived from the largest and most recent GWAS, which included data from 5.4 million individuals and identified over 12,000 genetic variants associated with variation in human height^8^. In this study, we used GWAS summary statistics of European including UK Biobank or without UK biobank (https://portals.broadinstitute.org/collaboration/giant/index.php/GIANT_consortium_data_files).

### Statistical analyses

#### PGS Development and Prediction without PCs

Standing height in UKB was residualized by regression over age, sex and 40 PCs, outliers (residualized height larger than 4) were removed, and residualized height was standardized.

We divided the European subset of the UK Biobank cohort into 90% for training (n = 367,060) and 10% for validation (n = 40,784). We validated the PGS in HCHS/SOL as well (n = 11,858) (Supplementary Fig. 1). The training cohort was used to estimate SNP effects, and the weights (coefficients) obtained were then applied to the validation cohorts. We fit the penalized regression model with “glmnet” function in Matlab where dependent variable is the standardized phenotype and independent variable is N x M matrix of standardized genotypes to obtain weights for each SNP (N: number of participants; M: number of SNPs). We selected SNPs for the model from publicly available GWAS summary statistics for standing height in European populations, including datasets with and without the UK Biobank.^25^ Specifically, when the training and validation was performed in UK Biobank prediction, we used GWAS of European data not including UK Biobank to avoid overfitting. When the training was performed in UK Biobank and prediction was performed in HCHS/SOL we used the European ancestry GWAS that included UK Biobank to increase sample size of the training cohort (Supplementary Fig. 1A). Given that penalized regression models can be sensitive to missing data, we included only those SNPs that were available across GWAS summary statistics, the UK Biobank cohort, and the HCHS/SOL cohorts. Ambiguous SNPs were excluded from the analyses. We assessed models using a range of p-value: 1 x 10^-16^, 1 x 10^-12^, 1 x 10^-8^, 1 x 10^-4^, 1 x 10^-3^, 1 x 10^-2^,1 x 10^-1^. The number of SNPs used to construct Polygenic Risk Scores (PGS) under different thresholds is summarized in Supplementary Table 1.

When fitting the penalized regression model, we set the alpha parameter of the glmnet R package to 1 for Lasso regression and 0 for Ridge regression. The model used 10 lambda values, resulting in 10 sets of SNP weights. We constructed 10 PGSs, one for each set of SNP weights (effect sizes), for individuals in the UK Biobank validation cohort (n = 40,784) and the HCHS/SOL cohort (n = 11,858) (Supplement Fig. 1B) by multiplying the number of risk alleles for each SNP by its corresponding weight and summing these weighted scores across all SNPs (algorithm 1). The 10 sets of PGS were then correlated with standardized phenotypes, and the squared correlation (R^2^) was estimated for the prediction accuracy. The PGS that performed best (with largest R^2^) was used in the analyses.

|  |
| --- |
| $PGS = \sum_{j=1}^M \beta_j * g_j$ (algorithm 1) |
| Where M is number of SNPs; $\beta_j$ is the weight of jth SNP effect allele; $g_j$ is the effect allele count of the jth SNP. |

#### PGS prediction with PCs

To estimate the PGS and PCs prediction accuracy, height was first residualized over age and sex, outliers (residualized height larger than 4) were removed and residualized height was standardized.

We randomly split the UK Biobank validation cohort (n = 40,784) and the HCHS/SOL cohort (n = 11,858) into 80% for training and 20% for validation (Supplementary Fig. 1C). The training data was used to fit linear regression models, with standardized height as the outcome variable. Each model included PGS and a varying number of PCs as predictors. This process was conducted separately for the UK Biobank European cohort (n = 32,627) and the HCHS/SOL cohort (n = 9,486).

The coefficients of PGS and PCs from the linear model were then applied to the 20% validation cohorts: UK Biobank European (n = 8,106) and HCHS/SOL (n = 2,372) to predict height (algorithm 2, using the top 5 PCs as an example). Prediction accuracy was assessed by the squared correlation between the predicted and standardized heights. Due to the limited sample size, we performed 1000 iterations of the 80/20 random split to ensure robustness and reliability of the results. The final metric is the average R^2^ over these iterations.

|  |
| --- |
| $Y = \beta_1 * PGS + \beta_2 * PC1 + \beta_3 * PC2 + \beta_4 * PC3 + \beta_5 * PC4 + \beta_6 * PC5$ (algorithm 2) |
| $\beta_1$ is the coefficient of PGS, $\beta_2$ to $\beta_6$ are the coefficients of top 5 PCs respectively. Y is the predicted standardized sex and age-adjusted height. |

Since the HCHS/SOL study includes multiple family members, there is a risk that relatives appearing in both the training and validation cohorts could artificially inflate the prediction accuracy. To mitigate this risk, we ensured that individuals with third-degree or closer familial relationships, as identified by kinship values greater than 0.0442, were included only in the testing group. We also applied the PGS trained from UKB EUR to different predominant ancestry subsets of HCHS/SOL (Supplementary Fig 3).

## Data Availability

All data produced in the present study are available upon reasonable request to the authors

## Supplementary

**Supplementary Fig 1.**
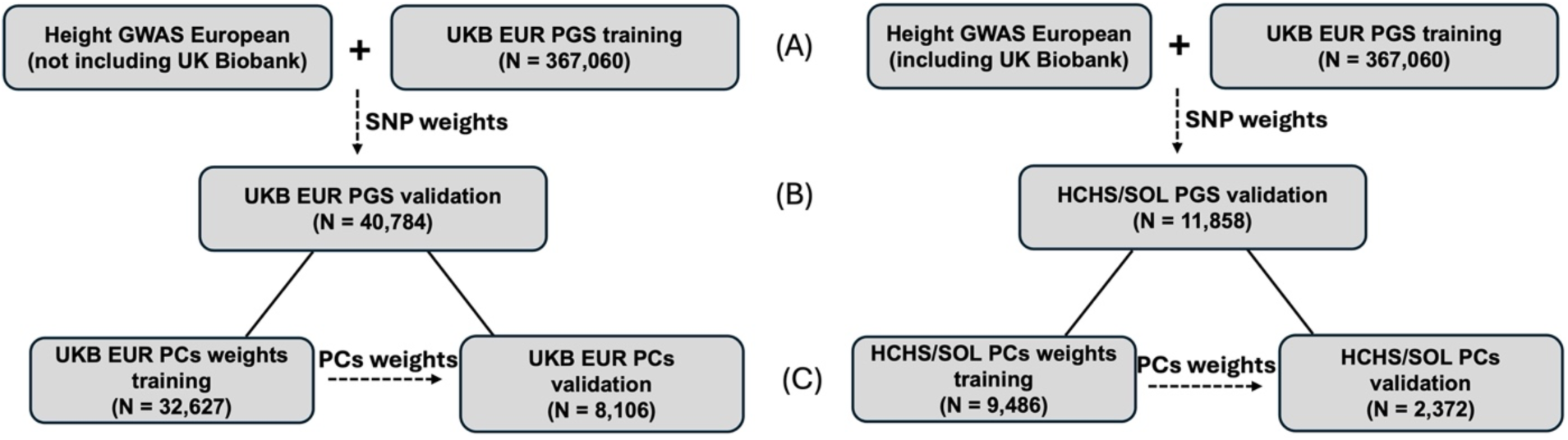
Analytic diagram in the main analyses and sample size used to train and test height prediction in UKB EUR and HCHS/SOL

**Supplementary Fig. 2.**
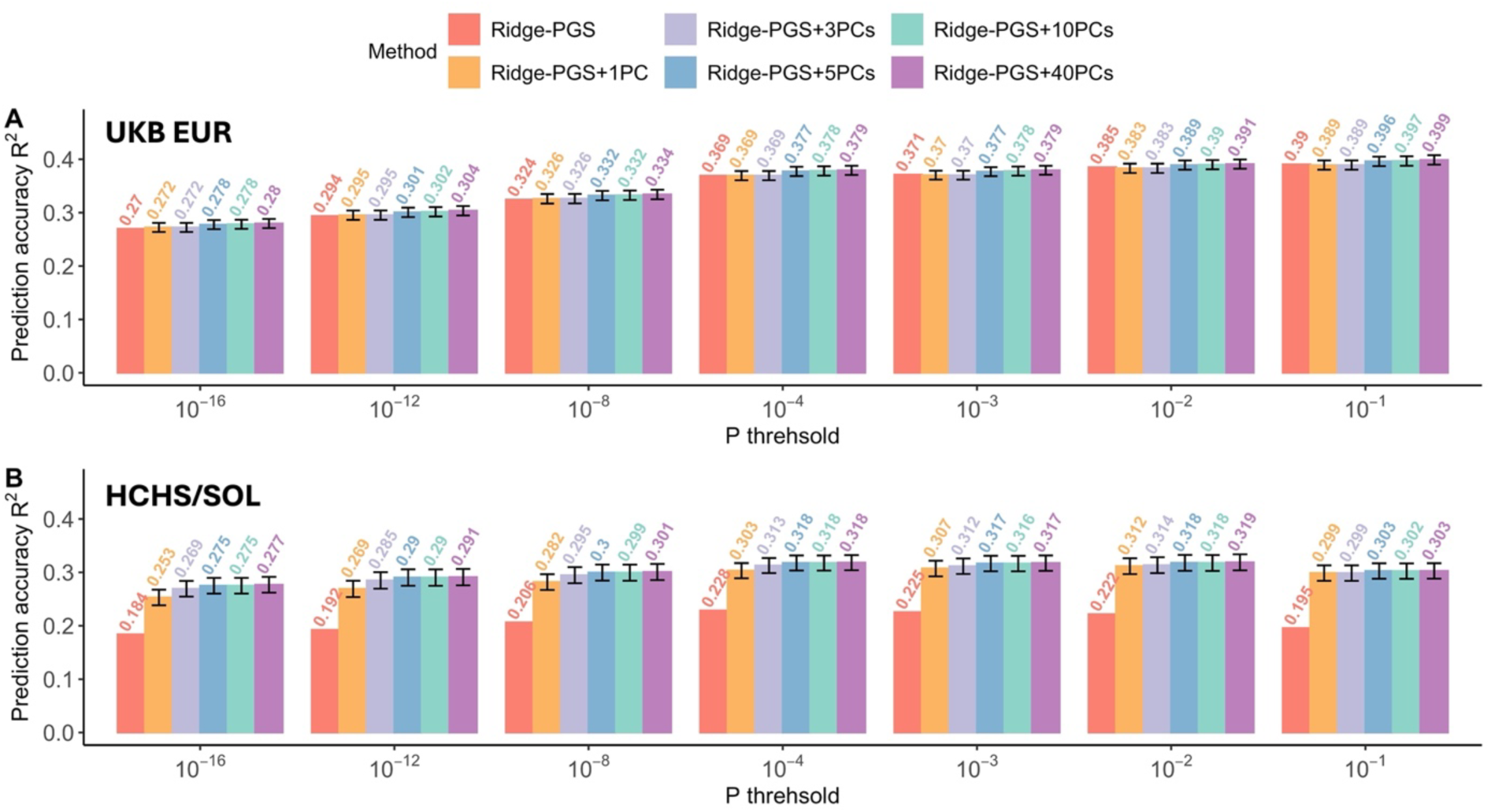
Predictive performance comparison on height between PGS only and combination of PGS and PCs using Ridge regression among UKB EUR and HCHS/SOL

**Supplementary Fig 3.**
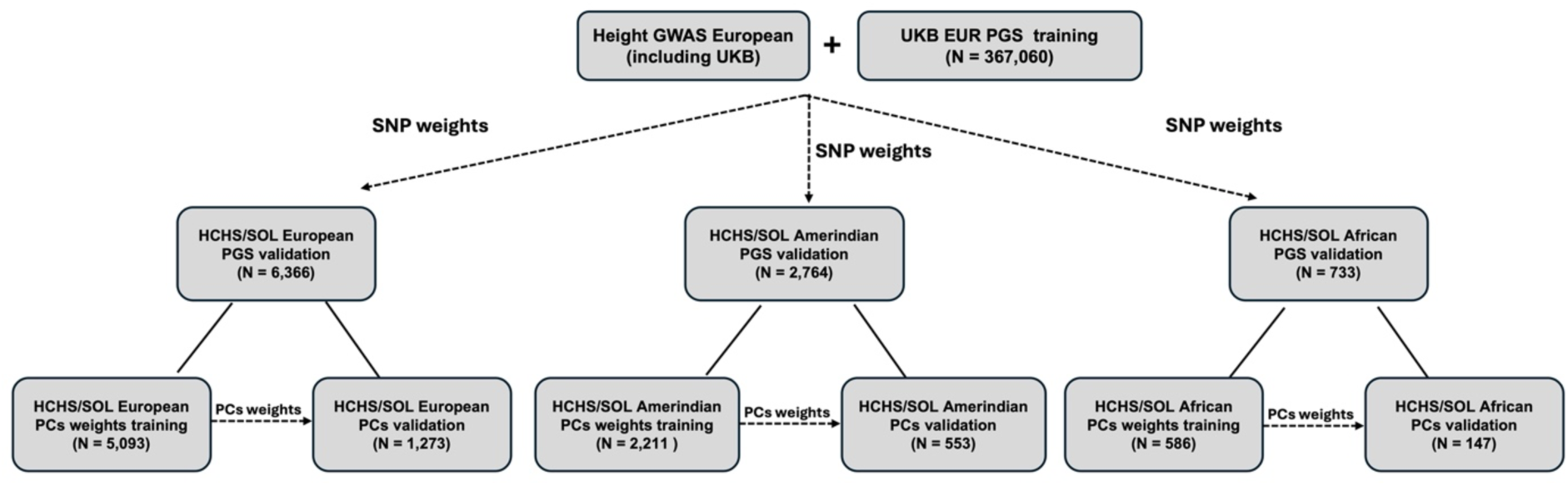
Analytic diagram and sample size used to train and test height prediction using UKB EUR-trained PGS in different ancestry subsets of HCHS/SOL

**Supplementary Fig. 4.**
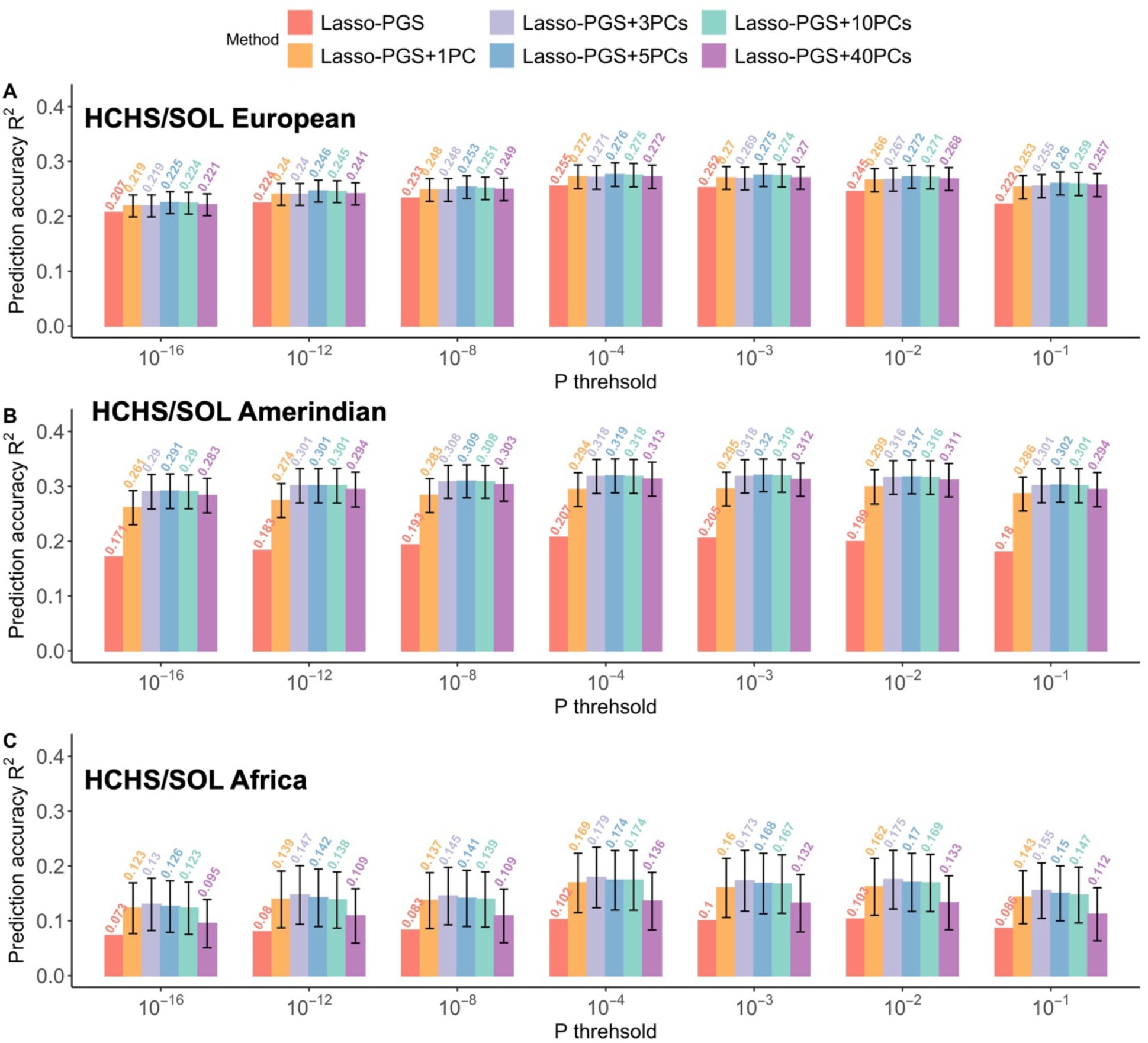
Predictive performance comparison on height between UKB EUR-trained PGS only and combination of PGS and PCs using Lasso regression in different ancestry subsets of HCHS/SOL European (A), American (B) and African (C) in HCHS/SOL.

**Supplementary Fig. 5.**
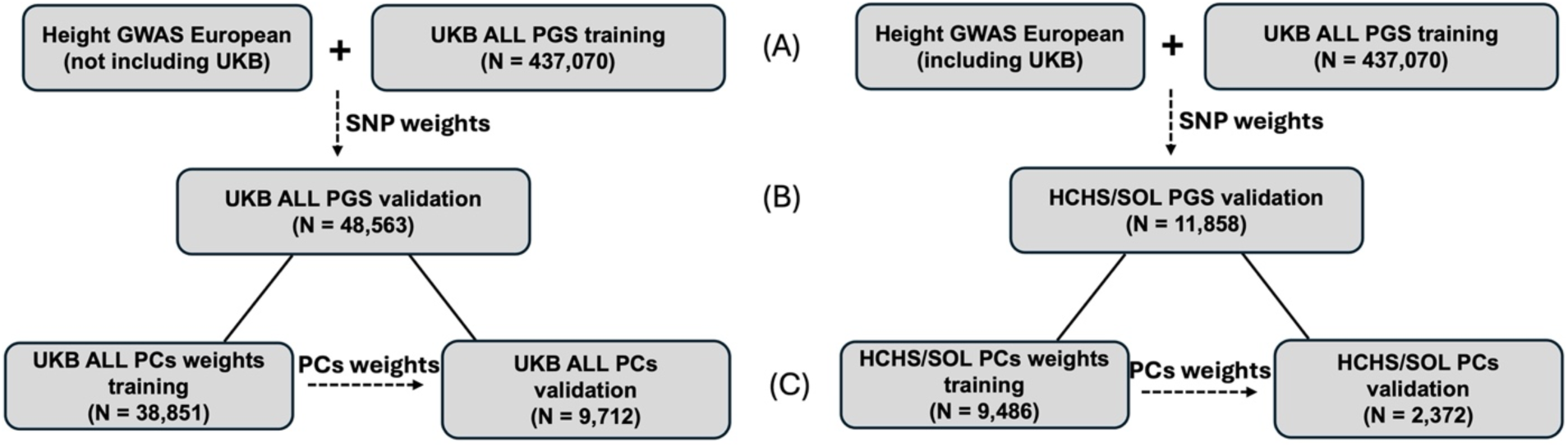
Analytic diagram in the main analyses and sample size used to train and test height prediction in UKB ALL and HCHS/SOL

**Supplementary Fig 6.**
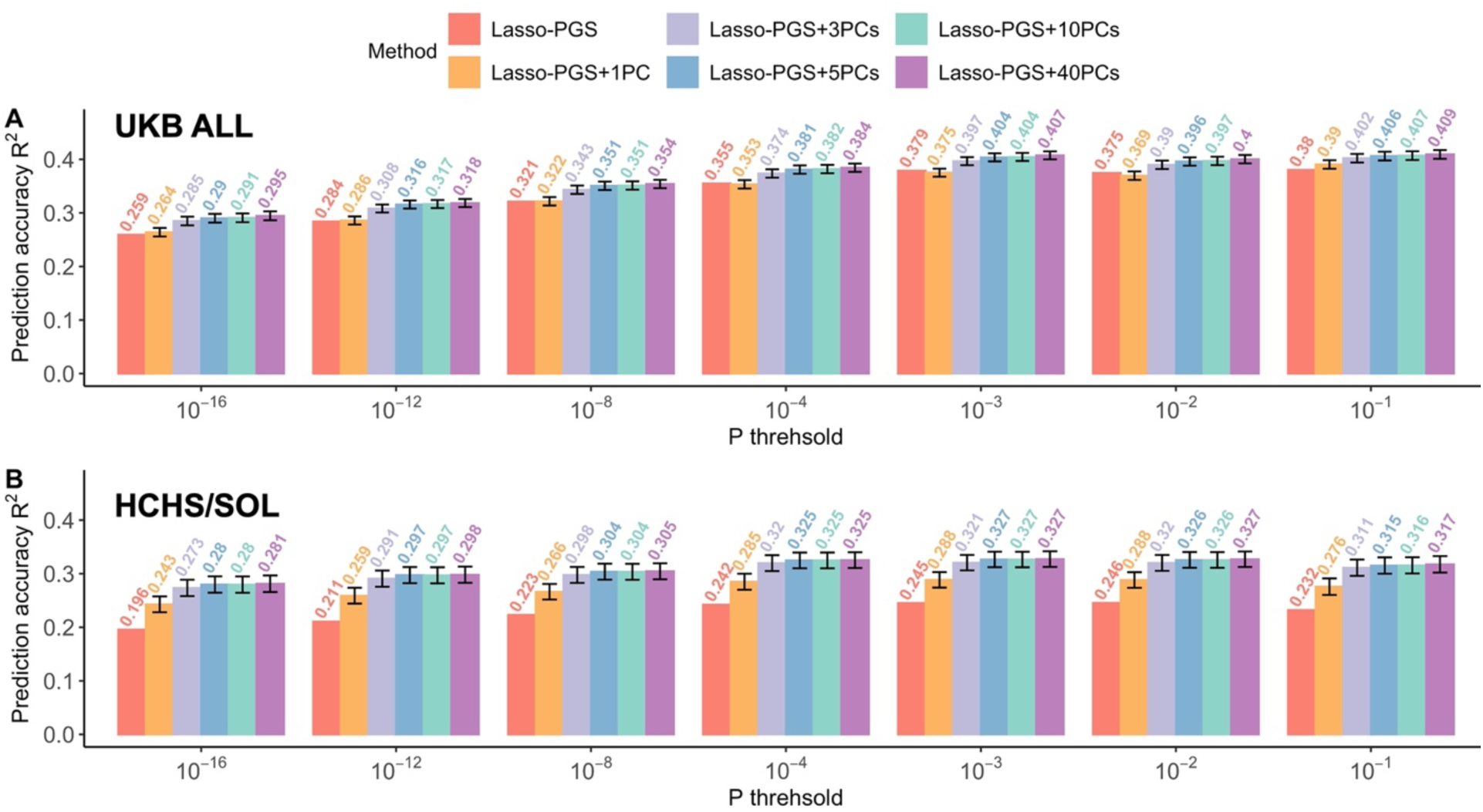
Predictive performance comparison on height between UKB ALL-trained PGS only and combination of PGS and PCs using Lasso regression among UKB ALL and HCHS/SOL

**Supplementary Fig. 7.**
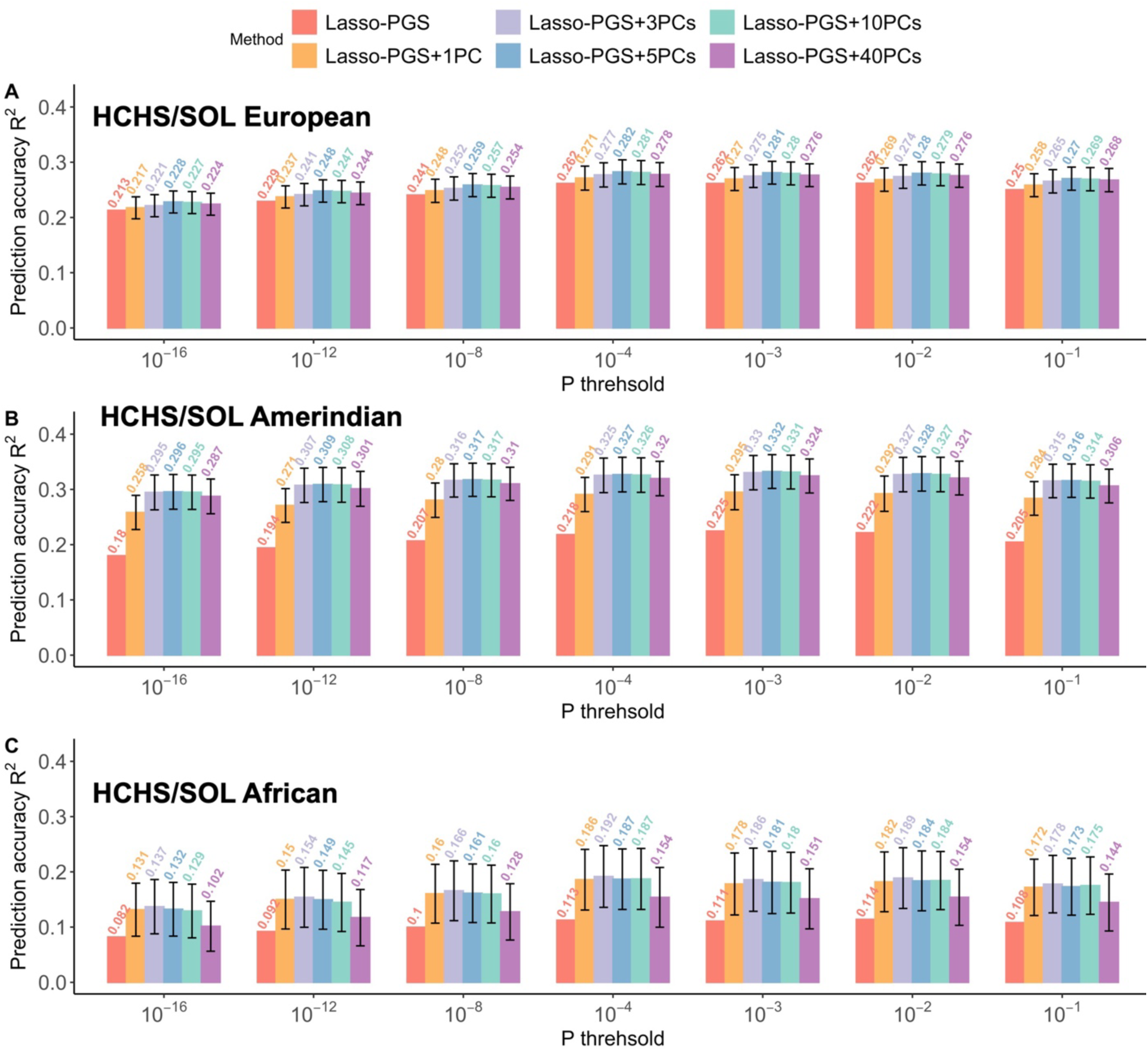
Predictive performance comparison on height between UKB ALL-trained PGS only and combination of PGS and PCs using Lasso regression in different ancestry subsets of HCHS/SOL European (A), American (B) and African (C) in SOLINCA

**Supplementary Table 1.** Number of SNPs in the prediction with different P threshold. No pruning was performed.

| GWAS not including UKB |  | GWAS including UKB |  |
| --- | --- | --- | --- |
| P threshold | N SNPs | P threshold | N SNPs |
| $< 10^{-16}$ | 21,383 | $< 10^{-16}$ | 39,411 |
| $< 10^{-12}$ | 30,649 | $< 10^{-12}$ | 53,669 |
| $< 10^{-8}$ | 49,556 | $< 10^{-8}$ | 80,738 |
| $< 10^{-4}$ | 105,394 | $< 10^{-4}$ | 154,159 |
| $< 10^{-3}$ | 141,666 | $< 10^{-3}$ | 197,606 |
| $< 10^{-2}$ | 210,670 | $< 10^{-2}$ | 272,654 |
| $< 10^{-1}$ | 372,205 | $< 10^{-1}$ | 432,066 |

